# A multipurpose machine learning approach to predict COVID-19 negative prognosis in São Paulo, Brazil

**DOI:** 10.1101/2020.08.26.20182584

**Authors:** Fernando Timoteo Fernandes, Tiago Almeida de Oliveira, Cristiane Esteves Teixeira, Andre Filipe de Moraes Batista, Gabriel Dalla Costa, Alexandre Dias Porto Chiavegatto Filho

## Abstract

**Introduction:** The new coronavirus disease (COVID-19) is a challenge for clinical decision-making and the effective allocation of healthcare resources. An accurate prognostic assessment is necessary to improve survival of patients, especially in developing countries. This study proposes to predict the risk of developing critical conditions in COVID-19 patients by training multipurpose algorithms.

**Methods:** A total of 1,040 patients with a positive RT-PCR diagnosis for COVID-19 from a large hospital from São Paulo, Brazil, were followed from March to June 2020, of which 288 (28%) presented a severe prognosis, i.e. Intensive Care Unit (ICU) admission, use of mechanical ventilation or death. Routinely-collected laboratory, clinical and demographic data was used to train five machine learning algorithms (artificial neural networks, extra trees, random forests, catboost, and extreme gradient boosting). A random sample of 70% of patients was used to train the algorithms and 30% were left for performance assessment, simulating new unseen data. In order to assess if the algorithms could capture general severe prognostic patterns, each model was trained by combining two out of three outcomes to predict the other.

**Results:** All algorithms presented very high predictive performance (average AUROC of 0.92, sensitivity of 0.92, and specificity of 0.82). The three most important variables for the multipurpose algorithms were ratio of lymphocyte per C-reactive protein, C-reactive protein and Braden Scale.

**Conclusion:** The results highlight the possibility that machine learning algorithms are able to predict unspecific negative COVID-19 outcomes from routinely-collected data.

## Introduction

The COVID-19 epidemic that first appeared in Wuhan, China, has rapidly spread worldwide and still continues to affect most countries, through late initial infections^12^ and second waves^34^. Currently, there have been more than 14 million cases and five hundred and ninety thousand confirmed deaths^5^. In critical patients, the disease has been shown to rapidly worsen a few days after infection^6,7^ requiring immediate clinical decisions, especially in developing countries with limited resources^8,9^. An accurate COVID-19 prognosis assessment is crucial for screening and treatment procedures and may increase patient survival^10,11^.

The consequences of COVID-19 have been disastrous for health systems in middle and low-income countries (LMICs) ^12^, especially in Brazil^13^. The lack of established knowledge about the disease has made it difficult to identify risk criteria to support clinical conducts and to allocate human and physical resources in health facilities and hospitals^14^. Currently, many Brazilian cities are at their saturation capacity for the provision of clinical care, especially regarding ICU beds and mechanical ventilators^15,16^.

Previous studies have used blood tests^17^, CT images^18,19^, sociodemographic and comorbidities history^20^ to develop COVID-19 diagnostic and prognostic models, including machine learning techniques^21-23^. Biomarkers from blood tests have emerged as important variables for poor prognostic factors^24^, which are a promising tool in poorer regions, due to its low cost and inclusion in standard protocols for clinical care. However, the majority of studies^25^ rely on algorithms trained on a single prognostic outcome, which in theory require the training of specific algorithms for each distinct negative outcome.

This study proposes to develop multipurpose machine learning algorithms to analyze if it is possible to predict overall poor prognosis for COVID-19 patients. We aim to test if the algorithms can generalize risk patterns for severe conditions, so they can be used as tools to assist in the prognosis of distinct negative outcomes for COVID-19 patients.

## Methods

### Data Source

A cohort of 3,280 patients with a RT-PCR diagnostic exam for COVID-19 from a large hospital chain in the city of São Paulo (BP-A Beneficência Portuguesa de São Paulo) were followed between March 1, 2020, and 28 June, 2020. Of these, 1,040 (31.7%) patients were positive for COVID-19 and were included in the analysis. The study was approved by the Institutional Review Board (IRB) of BP - A Beneficência Portuguesa de São Paulo (CAAE:31177220.4.3001.5421), including a waiver of informed consent. The study followed the guidelines of the transparent reporting of a multivariable prediction model for individual prognosis or diagnosis (TRIPOD).^26^

Individual patient data was collected from electronic medical records. We included as predictors only variables collected in early hospital admission, i.e. within 24 hours before and 24 hours after the RT-PCR exam. A total of 57 routinely-collected variables were used for the development of the predictive models, including demographic data, laboratory tests and vital signs (the complete list is described in Supplementary Table 1). Figure 1 illustrates the overall process.

**Fig. 1.**
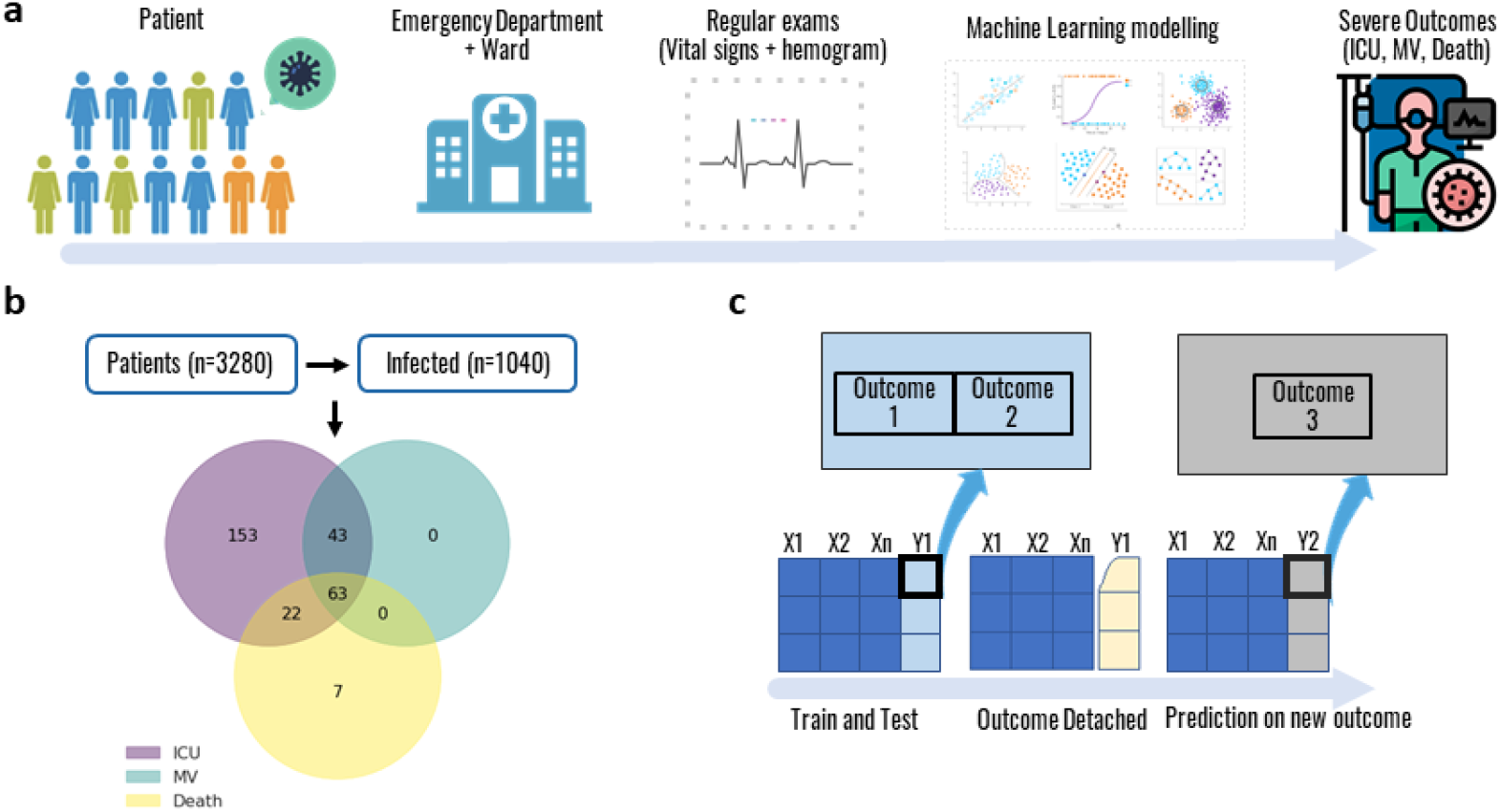
Process overview. **a** From hospital admission to a final outcome. **b** Population inclusion criteria and outcomes intersection. **c** The algorithm trained and tested using a combination of two outcomes. The same algorithm was then used to predict the remaining outcome.

### Machine learning techniques

Five of the most popular machine learning models for structured data (artificial neural networks^27^, extra trees^28^, random forests^29^, catboost^30^, and extreme gradient boosting^31^) were trained with 70% of the data, and tested in the other 30%, simulating new unknown data. All the results reported in this study are from the test set. K-fold cross-validation with 10 folds was used to adjust the hyperparameters with Bayesian optimization (HyperOpt). Due to the unbalanced nature of the outcomes, random undersampling was performed in the training set, by randomly selecting examples from the majority class for exclusion. This technique was implemented using the RandomUnderSampler imbalanced-learn class^32^.

Variables with more than two categories were represented by a set of dummy variables, with one variable for each category. Continuous variables were standardized using the z-score. Variables with a correlation greater than 0.90 (mean arterial pressure, total bilirubin, and creatine kinase) were discarded, and missing values were imputed by the median. To assess the performance of the models, measures such as accuracy, sensitivity (also known as recall), specificity, positive predictive value (PPV) (also known as precision), negative predictive value (NPV), and F1 score were analyzed. The value of the AUROC was used to select the best model.

To understand the individual contribution of each variable to the predictive models, we calculated their respective Shapley values. All the analyzes were performed using the Python programming language with the scikit-learn library.

### Patient and public involvement

Patients and the public were not directly involved in the design and conduct of this research.

## Results

### Descriptive statistics

Table 1 shows the descriptive statistics for the demographic characteristics of the patients. The sample of the study (1,040 patients with COVID-19) was mostly comprised by men (53.3%), with an average age of 51.7 years, and the majority of patients (63.8%) were white. The full descriptive statistics for all variables are presented in Supplementary Table 1.

**Table 1.** Descriptive statistics of the demographics characteristics of the sample.

| **Variable** | **ICU** | **MV** | **Death** | **Total** |
| --- | --- | --- | --- | --- |
|  | **Mean (SD)** | **Mean (SD)** | **Mean (SD)** | **Mean (SD)** |
| Age (years) | 63.2 (17.1) | 65.4 (16.2) | 73.7 (14.4) | 51.7 (18.9) |
| Weight (kg) | 79.80 (17.8) | 78.9 (16.4) | 74.5 (12.0) | 80.9 (18.7) |
| BMI | 28.2 (5.4) | 27.8 (4.4) | 27.1 (4.1) | 28.8 (5.9) |
| Height (cm) | 146.1 (56.5) | 147.9 (55.9) | 152.4 (47.1) | 154.9 (44.0) |
| Sex |  |  |  |  |
| Female (%) | 42.0 | 34.9 | 42.4 | 46.7 |
| Race (%) |  |  |  |  |
| Asian | 1.4 | 0.9 | 1.1 | 1.2 |
| White | 71.2 | 72.6 | 81.5 | 63.8 |
| Indigenous | 0.4 | 0.9 | 1.1 | 0.2 |
| Black | 3.6 | 1.9 | 1.1 | 3.2 |
| Mixed | 16 | 20.8 | 13 | 14.1 |
| N/A | 7.5 | 2.8 | 2.2 | 17.5 |

### Algorithms performance

We analyzed the predictive performance of the algorithms for three negative prognostic outcomes: ICU admission (n=263, 25.5%), mechanical ventilation (MV) intubation (n=106, 10.2%) and death (n=92, 9.4%). We first tested the predictive performance of the machine learning algorithms for a specific individual outcome (e.g. death). We then used the other two outcomes (in this specific example, mechanical ventilation and ICU admission) to train another model, and then tested this model performance to predict the previous outcome (death). We then compared the performance of the two strategies using the 95% confidence interval of the area under the receiver operating characteristic curve (AUROC).

Table 2 shows the results of the models trained with the aggregated outcomes and the models with a single outcome. Every model, even the ones trained with different outcomes, presented high predictive performance, always with an AUROC over 0.91 in the test set. The individual models were overall better, but the difference between the aggregated and individual models were all within the 95% confidence intervals. Supplementary Figure 1 shows the AUROC for each model. The sensitivity and specificity of the machine learning algorithms were also very high, in most cases over 0.8, with an average sensitivity of 0.92 and specificity of 0.82. In Supplementary Table 2 we present the final hyperparameters for each model.

**Table 2.** Predictive performance comparison in the test set for aggregated and individual models.

| Combination | Best algorithm | AUC [95% C.I.] | Sensitivity | Specificity | PPV | NPV | F1 |
| --- | --- | --- | --- | --- | --- | --- | --- |
| ICU + MV |  |  |  |  |  |  |  |
| *predict ICU* | Random Forest | 0.959 [0.94; 0.98] | 0.906 | 0.868 | 0.720 | 0.961 | 0.802 |
| *predict MV* |  | 0.912 [0.87; 0.95] | 0.935 | 0.723 | 0.271 | 0.990 | 0.420 |
| *predict Death* |  | 0.925 [0.89; 0.96] | 0.969 | 0.730 | 0.290 | 0.995 | 0.446 |
| Only Death |  |  |  |  |  |  |  |
| *predict Death* | Extra Trees | 0.972 [0.95; 1.00] | 0.964 | 0.863 | 0.409 | 0.996 | 0.574 |
| ICU + Death |  |  |  |  |  |  |  |
| *predict ICU* | XGBoost | 0.965 [0.95; 0.98] | 0.847 | 0.930 | 0.818 | 0.942 | 0.832 |
| *predict MV* |  | 0.925 [0.89; 0.96] | 0.946 | 0.808 | 0.398 | 0.991 | 0.560 |
| *predict Death* |  | 0.922 [0.89; 0.95] | 1.000 | 0.787 | 0.307 | 1.000 | 0.470 |
| Only MV |  |  |  |  |  |  |  |
| *predict MV* | Extra Trees | 0.945 [0.91; 0.98] | 0.906 | 0.819 | 0.362 | 0.987 | 0.518 |
| MV + Death |  |  |  |  |  |  |  |
| *predict ICU* | Random Forest | 0.921 [0.89; 0.95] | 0.765 | 0.901 | 0.729 | 0.917 | 0.747 |
| *predict MV* |  | 0.940 [0.91; 0.97] | 0.933 | 0.799 | 0.329 | 0.991 | 0.487 |
| *predict Death* |  | 0.943 [0.91; 0.98] | 0.963 | 0.794 | 0.306 | 0.996 | 0.464 |
| Only ICU |  |  |  |  |  |  |  |
| *predict ICU* | Random Forest | 0.959 [0.94; 0.98] | 0.906 | 0.868 | 0.720 | 0.961 | 0.802 |

### Interpretability

Figure 2 presents the prediction density for each individual outcome according to the different training strategies.

**Fig. 2.**
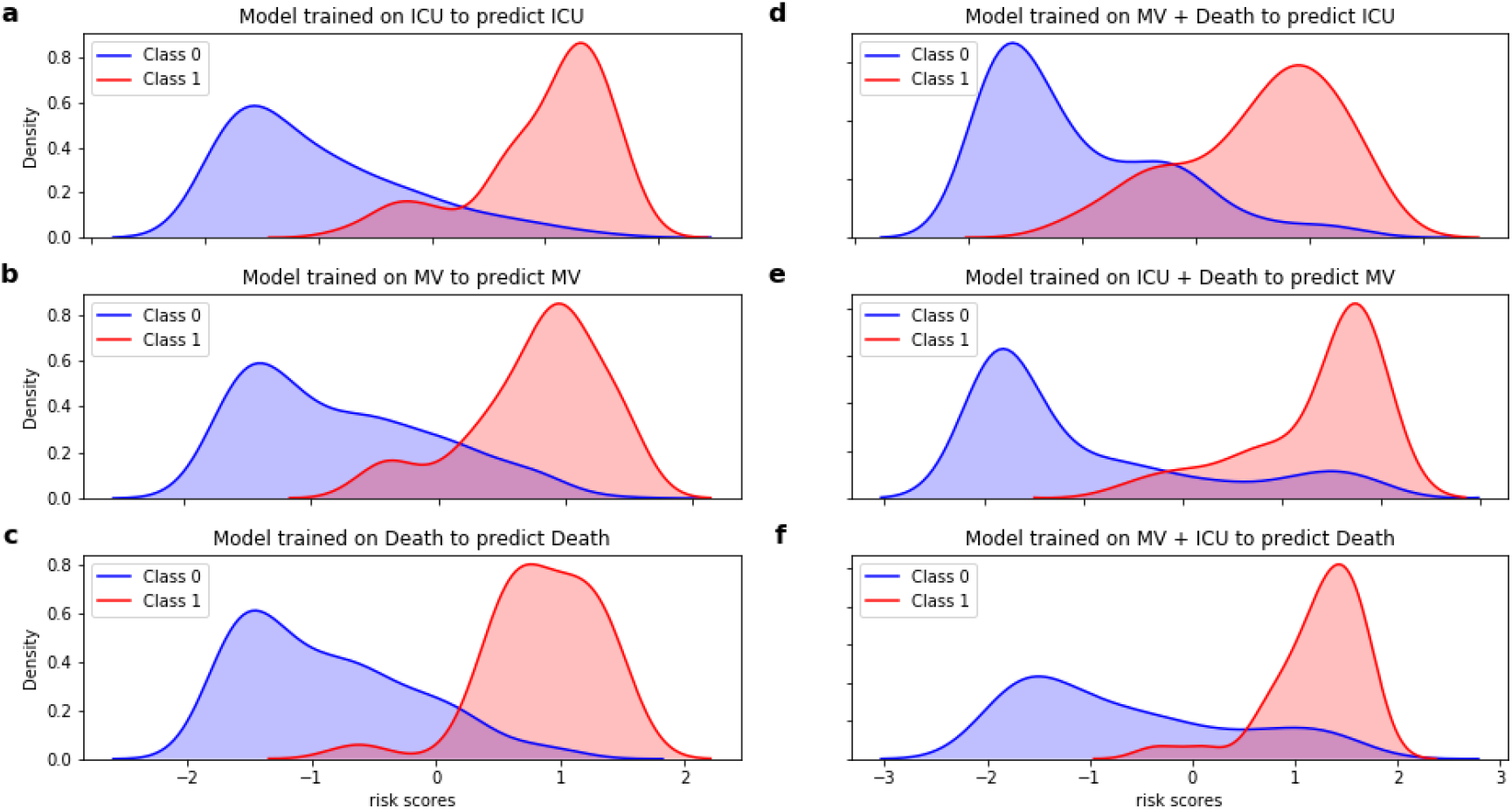
Density Plots. a-c. Density plots for single outcome models. d-f. Density plots for aggregate models predicting unspecific outcome.

The results point to a low overlap between negative and positive cases, indicating a good discriminative ability of the algorithms irrespective of the training strategy.

Figure 3 presents the top 5 variables that most contributed to prediction in the aggregated models, according to the Shapley values.

**Fig. 3.**
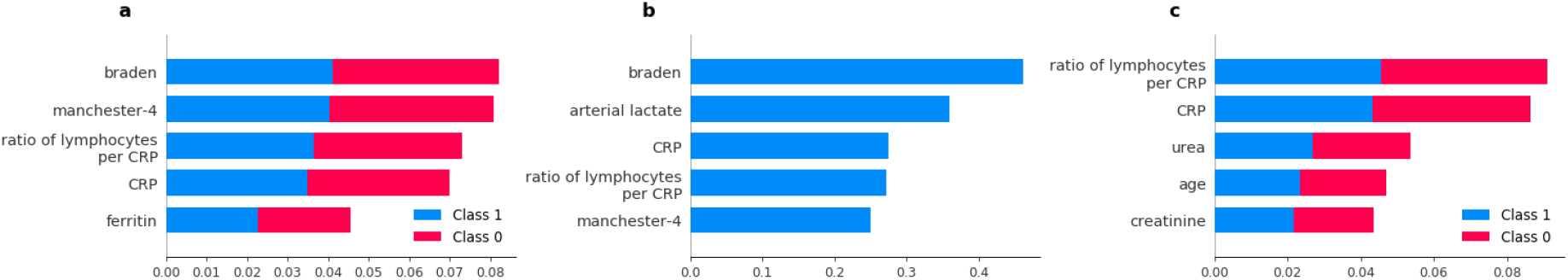
Top five feature contributions to the predictive models, according to Shapley Values. a. MV+ICU b. Death + ICU c. Death + MV

The Braden score played an important role in the aggregated outcome algorithms, ranking as the most important predictor in two of three models. Also, the C-reactive protein and ratio of lymphocytes per C-reactive protein proved to be good predictors, appearing in the top 5 in all three models. Urea, age, creatinine, and arterial lactate were important for only one of the aggregated models.

## Discussion

We found that machine learning algorithms were able to predict with high overall performance negative prognostic outcomes for COVID-19, even when the specific outcome was not included in the training of the algorithms. All models presented an AUROC higher than 0.91 (average of 0.92) in the test set, with high sensitivity and specificity (average of 0.92 and 0.82, respectively). The results highlight the possibility that high-performance machine learning algorithms are able to predict unspecific negative COVID-19 outcomes using routinely-collected data.

Brazil is currently the second country in the world in total number of cases and deaths from COVID-19^33^. There is a growing demand in Brazil, and in many other developing countries, for decision support in the allocation of scarce hospital resources, especially in relation to the availability of ICU beds and mechanical ventilators^34,35^. From a clinical standard, knowledge about immediate risks of negative prognosis can also contribute to the early start of preventive measures and new interventions, and thereby increase patient survival^10,11^.

The results of this study highlight that it is possible to predict with high performance the risk of a negative prognosis in patients with COVID-19. Additionally, we found that it is also possible to predict the risk of negative outcomes well even when they were not used for training of the model. This result is promising for the prediction of specific outcomes that are not possible to be collected for model training, either due to technical difficulties in registering the presence of these outcomes, or in the case of predicting a new negative outcome previously unknown.

The development of multipurpose prognostic algorithms, i.e. algorithms that identify nonspecific outcomes and overall future clinical deterioration, can be used in a large number of situations, especially in the case of complex and unknown diseases that lead to the development of several different negative outcomes. Instead of having to develop a different algorithm for each of the specific outcomes, multipurpose models can provide more comprehensive and clinically relevant information about the risks of future health problems of patients.

For every outcome, variable importance analysis identified that age, C-reactive protein (CRP), creatinine, urea and the Braden Scale were usually among the most important. While the age of the patient is widely found to be an important predictor for most negative health outcomes, CRP has been increasingly included among the main inflammatory biomarkers for the prognosis of cardiovascular^36^ and respiratory diseases^37^. High levels of CRP have been also previously associated with individual severity of SARS-CoV-2^38,39^. Interestingly, previous studies have also identified that chronic kidney disease is associated with developing severe conditions in COVID-19 patients^40-42^, where it has been observed that patients with higher levels of creatinine and urea are more at risk^43^. The Braden Scale is often used as a predictor for pressure ulcers, a common clinical classification scale for predicting pneumonia^44^ during clinical reception, and in this study, it was an important predictor for negative prognosis in COVID-19 patients. The scale has a score between 1 (worst score) and 4 (best score) where the factors included are sensory perception, skin moisture, activity, mobility, nutritional status and friction^45^. The percentage of lymphocytes in the blood has been described as a strong predictor of prognosis for the severity of the new coronavirus. The randomized study by Lin Tan et al.^46^ suggests that, in most confirmed cases, the percentage of lymphocytes was reduced to 5% in two weeks after the onset of COVID-19, in line with other studies findings^47^.

The study has a few limitations that need to be mentioned. First, some of the outcomes overlap which may have helped the performance of the aggregated models, even though in the majority of cases the outcomes were independent. In the case of ICU admission, 55% of the patients did not die or used MV, while in the case of MV and death, 63% and 70% of their respective aggregated model was trained on other outcomes. Ideally, the outcomes would never overlap, but this is clinically unfeasible given the interlaced nature of negative prognostic outcomes. Another limitation is that we analyzed data from an urban COVID-19 hotspot in Brazil, in a period where clinical protocols for the disease were still being established, so this could affect the incidence of prognostic outcomes and may not directly generalize to other periods.

## Conclusion

In conclusion, we found that machine learning algorithms can predict severe outcomes in COVID-19 patients with high performance, including previously unobserved outcomes, using only routinely-collected laboratory, clinical and demographic data. The use of multipurpose algorithms for the prediction of overall negative prognosis is a promising new area that can support doctors with clinical and administrative decisions, especially regarding priorities for hospital admission and monitoring.

## Data Availability

The data comes from medical records from BP - A Beneficencia Portuguesa de Sao Paulo Hospital in Brazil and it is not publicly available as it contains sensitive information of patients.

## Data availability

The data comes from medical records from BP - A Beneficência Portuguesa de São Paulo Hospital in Brazil and it is not publicly available as it contains sensitive information of patients.

## Code availability

All the code written to process and analyze the data can be made available upon request to the corresponding author.

## Acknowledgements

We would like to thank the BP - A Beneficência Portuguesa de São Paulo Hospital for its willingness to contribute to the research. This work was supported by National Council for Scientific and Technological Development (CNPq) under Grant Number 402626/2020-6 and Paraíba Research Foundation FAPESQPB with Grant Number 206/2020.

## Author information

### Contributions

Initial study concept and design: A.D.P.C.F. Acquisition of data: G.D.C. Model training: F.T.F, T.A.O, C.E.T, A.F.M.B. Analysis and interpretation of data: F.T.F, T.A.O, C.E.T, G.D.C., A.D.P.C.F. Drafting of the paper: All authors contributed for drafting the manuscript. Critical revision of the manuscript: All authors provided critical review of the manuscript and approved the final draft for publication.

### Corresponding author

Correspondence should be adressed to Fernando T. Fernandes.

